# Vaccinating the Frontlines: A Qualitative Exploration of Hospital Healthcare Worker Perspectives on Influenza and COVID-19 Immunization

**DOI:** 10.1101/2024.07.10.24310248

**Authors:** Erica N. Rosser, Sabra L. Klein, Richard E. Rothman, Andrew Pekosz, Rosemary Morgan

## Abstract

**Introduction:** Although they face higher occupational risk of contracting viral respiratory infections, hospital healthcare worker vaccine hesitancy persists. While most studies have used survey methods to quantify the prevalence of and reasons for healthcare worker vaccine hesitancy, this study employs a qualitative approach to understand their attitudes and beliefs associated with influenza and COVID-19 vaccination.

**Methods:** To understand frontline healthcare worker experiences and perspectives on influenza and COVID-19 vaccination, 30 semi-structured interviews were conducted in summer/fall 2022 with staff recruited from two Johns Hopkins hospitals in Maryland. An in-depth, key informant interview was conducted with an expert in public health audience engagement. Interviews were audio recorded and transcribed for thematic and Framework analysis using NVivo software (QSR International, Melbourne, Australia).

**Results:** Healthcare workers engaged in little influenza vaccine information seeking due to their familiarity with the disease and low perceived disease severity. Approximately half (n=16) of healthcare workers reported no vaccine hesitancy towards influenza or COVID-19 vaccines. No physicians or physician assistants expressed any vaccine hesitancy, while most nurses expressed some (n=10). More than half of the women (n=14) expressed COVID-19 vaccine hesitancy compared to none of the men. Structural factors including hospital tier, unit assignment, and professional role influenced perceived risk of disease exposure and subsequent healthcare worker vaccination decisions. Institutional policies, including mandates and a pro-vaccine environment encouraged vaccination uptake. Healthcare workers reported being more receptive to vaccine messaging that focused on protection from disease, scientific and public health data and their heightened occupational exposure to pathogens.

**Conclusions:** Despite their medical knowledge, healthcare workers are susceptible to vaccine hesitancy. Strategies to address specific concerns are needed and can be informed by our findings. A flexible and multi-pronged approach that considers individual anxieties, workplace structures, and the need for open communication with tailored messaging is necessary to promote vaccine acceptance in healthcare settings.

**KEY MESSAGES:** *What is already known on this topic:* Healthcare worker vaccine hesitancy has been associated with many factors including race, gender, age and concerns about vaccine safety.

*What this study adds:* Much of the research on healthcare worker vaccine hesitancy has used surveys and questionnaires giving a broad description of the prevalence and patterns of vaccine hesitancy in the healthcare workforce. This qualitative study examines vaccine behavior (rather than merely intent) through a cross comparison of healthcare workers’ experiences and attitudes towards influenza and COVID-19 vaccination.

*How this study might affect research, practice or policy:* Study findings can be used to help tailor vaccine messaging to hospital healthcare workers which could offset concerns regarding vaccine efficacy and risk, to promote vaccine uptake.

## INTRODUCTION

Hospital healthcare workers (HCWs) at the forefront of healthcare delivery provide essential care to patients suffering from viral respiratory illnesses including influenza and COVID-19. They also have an increased risk of contracting and transmitting these viral respiratory illnesses to others they encounter (i.e., co-workers, patients and their own families)[1,2].Vaccination is one critical tool that protects hospital HCWs from illness (including severe illness), and others they come in contact with from transmission[3]. HCWs are frequently mandated to receive influenza vaccines for work and were prioritized for COVID-19 vaccination during the pandemic[4–6]. However, despite the clear benefits of vaccination, evidence shows that vaccine hesitancy, defined by the WHO as “a delay in acceptance or refusal of vaccination despite availability and accessibility of vaccine services,” persists among HCWs [7].

In December of 2020 when the COVID-19 vaccine roll-out was introduced, a national survey found that about 30% of people working in healthcare delivery settings in the US expressed COVID-19 vaccine hesitancy[8]. Prior studies reported that individual HCW decision-making around vaccines vary markedly and are driven by a multitude of psychosocial factors, as well as age, race, gender, income-level and specific role of the HCW in the hospital system[9,10]. Similar to the general population, HCWs may also harbor concerns about vaccine safety and efficacy, and may rely on misinformation or disinformation, or lack trust in healthcare authorities [11].

In recent years, there has been a growing focus on understanding and addressing vaccine hesitancy among frontline HCWs. However, much of the research to date has relied on surveys and questionnaires [9,12–14]. Qualitative studies can enhance understanding of the complex and nuanced perspectives that drive immunization decision-making and be used to inform targeted interventions. The present study explores the perspectives of frontline HCWs at two Maryland hospitals on their decisions about getting vaccinated against influenza and COVID-19. This qualitative study identifies motivations and concerns that contribute to individual HCWs’ vaccination decisions, providing valuable insights for addressing vaccine hesitancy and developing future methods to promote vaccination uptake among this priority population.

## MATERIALS AND METHODS

### Healthcare setting and vaccination policies

Johns Hopkins Health System (JHHS) maintains a comprehensive network of healthcare services across the Baltimore–Washington D.C metropolitan area including five hospitals, as well as centers, community practices and homecare [15]. Johns Hopkins Medicine (JHM) has a mandatory policy requiring all personnel – except those who qualify for a medical or religious exemption – to receive the annual influenza vaccine. In 2021, JHM announced a COVID-19 vaccination policy requiring all non-exempt personnel to be fully vaccinated by September 1, 2021 (Johns Hopkins Medicine*. Update on JHM COVID-19 Vaccination Requirement* [email]. 2021 01 July). In the summer and fall of 2022, two years after providers began caring for COVID-19 patients in their respective clinical settings, we interviewed adult frontline HCWs working in high-risk departments (defined by risk of exposure to COVID-19 positive patients) at two JHHS hospitals, the Johns Hopkins Hospital (JHH) in urban Baltimore City and Howard County General Hospital (HCGH) in Columbia, MD, a suburb of Baltimore.

### Participant Identification and Recruitment

Over several months, multiple methods were used to recruit study participants including posting flyers at JHH and HCGH and announcements in internal newsletters. We also recruited participants directly by attending HCW unit meetings (i.e., clinical “huddles”) and through snowball sampling once data collection began. Interviews were restricted to HCWs above the age of 18 whose work involved direct patient contact and who self-reported having received the influenza vaccine in the year preceding the start of the COVID-19 pandemic. From 52 inquiries, we screened HCWs to achieve data saturation, yielding a sample size (n=30) consistent with established norms for this type of research[16]. We also purposively recruited a Johns Hopkins School of Public Health staff member with more than a decade of experience working in audience engagement for expert opinion. This key informant was directly involved in crafting and disseminating public health messaging on influenza (pre- and post-COVID-19) and COVID-19 vaccination for both the Johns Hopkins community and the general public and was able to provide valuable insight into the development and delivery of vaccine acceptance strategies.

### Qualitative Data Collection and Analysis

Semi-structured interview guides were used for both the key informant and HCW interviews. All interviews were conducted via password-protected Zoom calls. Following informed consent, interviews were audio-recorded, and transcribed verbatim. Field notes written directly following interviews were appended to deidentified transcripts to provide additional context for analysis. Participants received a $50 gift card as a token of appreciation for their time.

The study codebook included *a priori* codes derived from the research objectives and emergent codes identified through transcript review, and study team debriefing and reflection. NVivo data analysis software (QSR International, Melbourne, Australia) was used to facilitate data management and organization including coding and production of code reports. Codes were aggregated into relevant themes, and memos synthesizing findings were created for each theme. Ethical approval was granted by the JHM Institutional Review Board (No. IRB00284889).

## RESULTS

### Summary of Data Collection

We conducted a total of 30 qualitative interviews with HCWs (**Table 1**) from JHH (19 interviews) and HCGH (11 interviews) with most participants being female, Caucasian, and nurses. About two-thirds of the HCWs across the two hospitals worked in the highest exposure units – emergency departments (ED), intensive care units (ICU) or COVID-19 units (n=21) – during the pandemic.

**Table 1.**
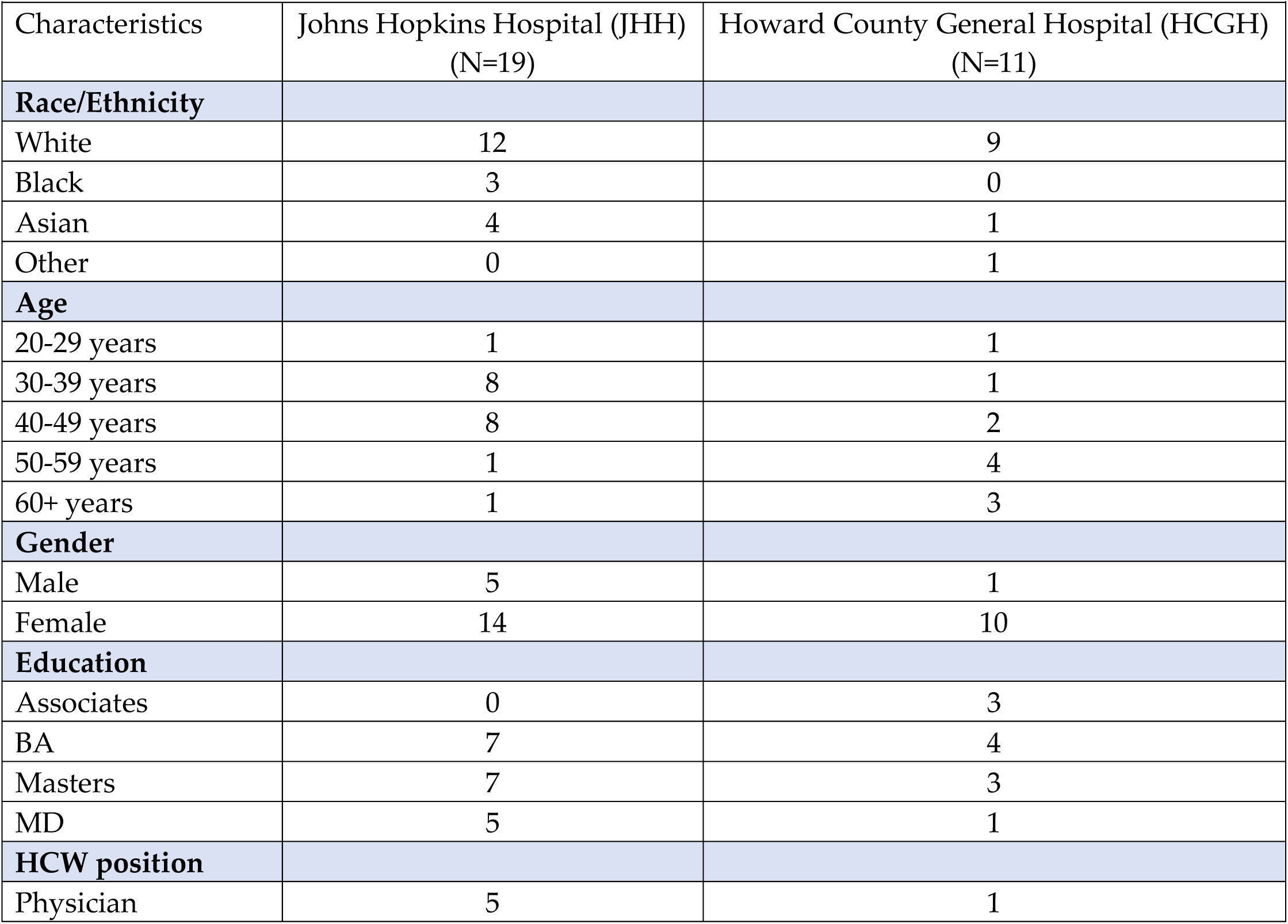
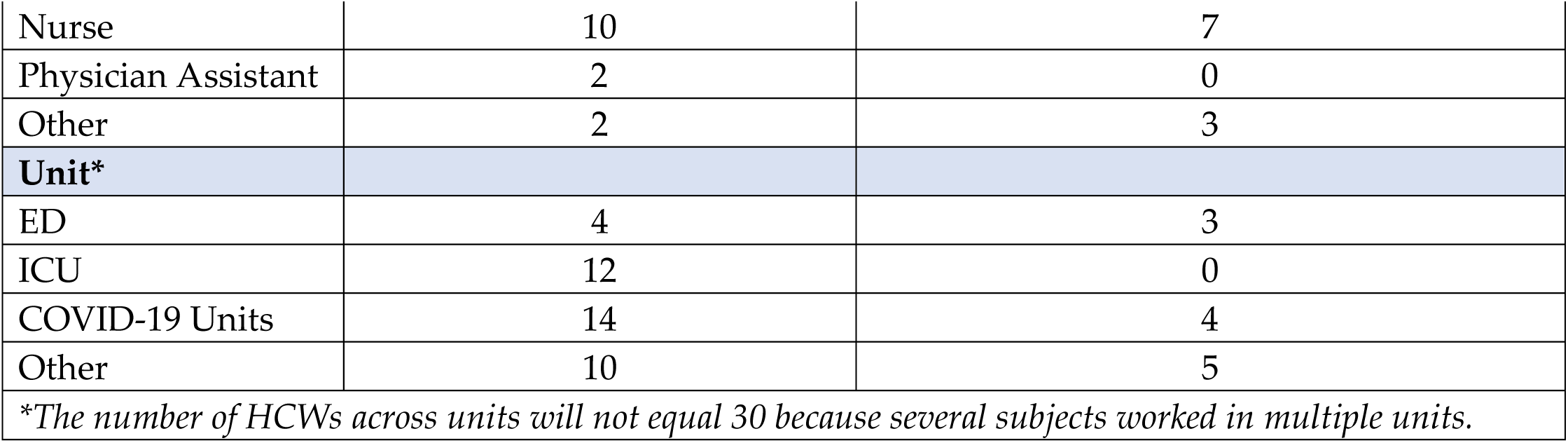
Interviewee Demographics.

| Characteristics | Johns Hopkins Hospital (JHH)<br>(N=19) | Howard County General Hospital (HCGH)<br>(N=11) |
| --- | --- | --- |
| **Race/Ethnicity** |  |  |
| White | 12 | 9 |
| Black | 3 | 0 |
| Asian | 4 | 1 |
| Other | 0 | 1 |
| **Age** |  |  |
| 20-29 years | 1 | 1 |
| 30-39 years | 8 | 1 |
| 40-49 years | 8 | 2 |
| 50-59 years | 1 | 4 |
| 60+ years | 1 | 3 |
| **Gender** |  |  |
| Male | 5 | 1 |
| Female | 14 | 10 |
| **Education** |  |  |
| Associates | 0 | 3 |
| BA | 7 | 4 |
| Masters | 7 | 3 |
| MD | 5 | 1 |
| **HCW position** |  |  |
| Physician | 5 | 1 |
| Nurse | 10 | 7 |
| Physician Assistant | 2 | 0 |
| Other | 2 | 3 |
| **Unit\*** |  |  |
| ED | 4 | 3 |
| ICU | 12 | 0 |
| COVID-19 Units | 14 | 4 |
| Other | 10 | 5 |
| *\*The number of HCWs across units will not equal 30 because several subjects worked in multiple units.* |  |  |

### Healthcare Workers Perceptions of Disease

Analysis uncovered differences in the frequency and depth of HCW’s discussion of influenza and COVID-19. When the subject of influenza emerged unprompted in interviews, it was primarily employed as a point of comparison with COVID-19.

#### Disease Severity and Susceptibility

HCWs across all units and hospital roles consistently reported minimal concern about occupational influenza exposure which they attributed to familiarity with influenza season and lower perceived disease severity. HCWs described influenza season as a predictable event with established infection control protocols, including increased hand hygiene and exercising droplet precautions. Compared to COVID-19, influenza was generally described as “*disagreeable and inconvenient*” rather than as a potentially life-threatening condition. This perception resulted in less information seeking and scrutiny surrounding the influenza vaccine, *“The flu vaccine is normal. We just get it every fall. I don’t think about it. It doesn’t matter what you email me or send me, I’m always going to get it.” (HCW09).* Influenza vaccine hesitancy was primarily limited to individuals with previous adverse reactions, rather than broader anxieties about the vaccine itself.

Conversely, most HCWs did express concern about contracting COVID-19. Those working in units with the highest disease rates early in the pandemic likened the experience to working in a “*hot zone*”. Fear of workplace transmission and being “*patient zero*” in their households led them to take rigorous precautions to protect their families (i.e., showering at work, using disposable bags for hospital attire). While most HCWs felt *“much safer”* about returning to normal activities and post-shift routines after vaccination, many said the decrease in worry predated vaccination and was more due to additional knowledge gained about key modes of COVID-19 virus transmission (i.e., close personal contact, breathing respiratory droplets). Several participants also reported that over time their anxieties shifted from concerns about patient-to-HCW transmission to concerns about potential transmission from unmasked colleagues in common areas such as staff break rooms where masking practices were perceived to be less stringent.

The few HCWs who reported minimal or no anxiety about COVID-19 infection during the pandemic attributed their lack of worry to their extensive experience working with infected populations – including some infected with pathogens they considered more dangerous than SARS-CoV2. One ED HCW explained, *“I wasn’t worried about catching COVID… A lot of my HIV-positive patients had C. diff, MRSA, tuberculosis, and I’m like, ‘I would rather get COVID than tuberculosis.’” (HCW04)*, illustrating their view of COVID-19 as another occupational risk that could be managed with proper training and infection control measures.

#### Impact

In terms of impact, the stress, hypersensitivity and emotional burden HCWs described experiencing over the course of the COVID-19 pandemic was much worse than during a typical influenza season in the same environment. HCWs attributed this disparity to the distinct characteristics of each illness. An ED physician described how influenza patients typically experience either rapid improvement or decline, *“whereas with COVID, it’s almost like a slow burn that just continues to burn hotter and hotter over the next two weeks… That slow decline is much more mentally scarring.” (HCW03).* An ICU physician corroborated the emotional toll of the unprecedented severity and scale of the COVID-19 surge compared to influenza season, *“I remember nights in the COVID ICU where all 24 beds, every single patient had COVID. Each person was prone, paralyzed, on maximal amounts of oxygen from the ventilator. That never would happen in a flu season. It’s comparing a whole hospital response in a major pandemic to… [something that] just seems less novel, less important, less dangerous.” (HCW27)*.

### Vaccine Hesitancy

Approximately half of HCWs expressed no hesitancy (n=16) towards either influenza or COVID-19 vaccination. Notably, no physicians or physician assistants (PAs) expressed any vaccine hesitancy, while most nurses expressed some towards either influenza or COVID-19 vaccination (n=10). Similar ratios of HCWs expressed vaccine hesitancy across the two participating hospitals.

#### Vaccine Hesitancy and Vaccination Timing

All participants who expressed no vaccine hesitancy (n=16) reported receiving their first COVID-19 vaccine early in the rollout between December 2020 and January 2021. Approximately half (n=8) of those who expressed some degree of vaccine hesitancy still reported having received their first dose within the same early timeframe. The remaining HCWs who expressed vaccine hesitancy delayed their vaccination until later in the year (**Figure 1**).

**Figure 1.**
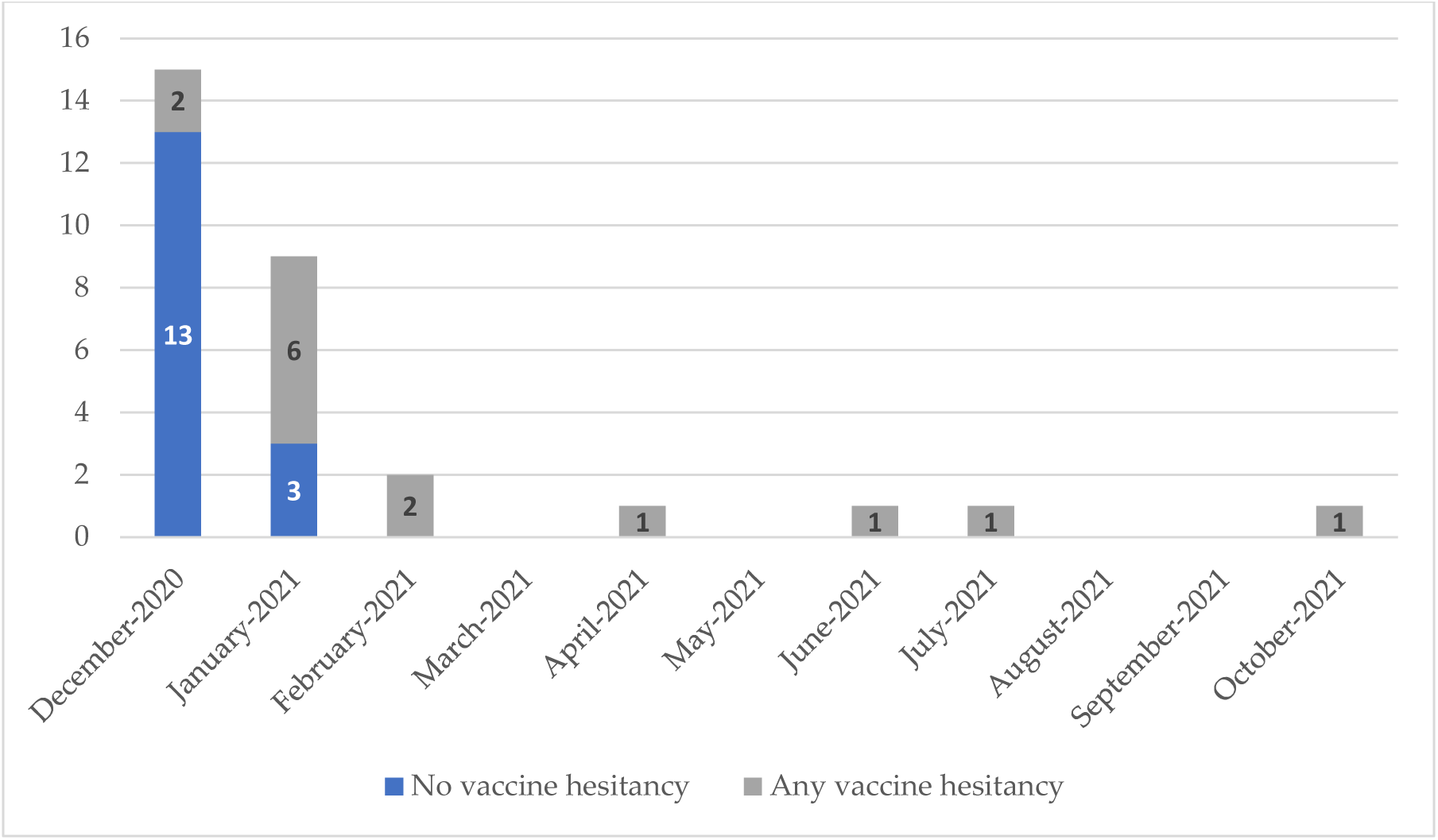
**Date of First HCW COVID-19 Vaccine by Vaccine Hesitancy Expressed**

#### Vaccine Hesitancy and Sociodemographic Characteristics

##### Gender and Vaccine Hesitancy

Analysis revealed distinct differences in COVID-19 vaccine hesitancy when comparing men and women. Interestingly, none of the men expressed any vaccine hesitancy towards either influenza or COVID-19 vaccines. Conversely, of the 24 women interviewed, more than half (n=14) expressed some degree of COVID-19 vaccine hesitancy (**Figure 2**). Female HCWs frequently discussed their concerns related to vaccine safety, as well as their age, gender, and reproductive health. Safety concerns often centered on the perceived rapid speed of the vaccine development process, while discussions surrounding gender focused on anxieties about potential reproductive impacts and the initial lack of clear information on this early in the pandemic. HCWs of both genders highlighted other factors influencing their decisions including the impact of their families’ views and health status and their individual health histories. They also discussed the impact of varied education levels and their training and knowledge of infection prevention.

**Figure 2.**
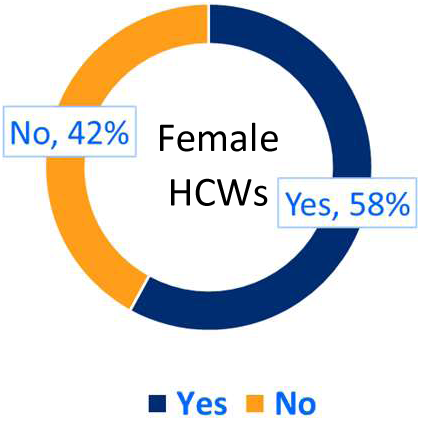
**Percentage of Female HCWs who Expressed any COVID-19 Vaccine Hesitancy**

##### Educational Level and COVID-19 Vaccine Hesitancy

While analysis found no instances of influenza or COVID-19 vaccine hesitancy among partcipants holding a MD degree (n=6), among the other (non-MD) participants, the rate of vaccine hesitancy was higher among those with higher educational training: 33% of those with an Associate degree (n=1), 63.6% of those with a Bachelor’s degree (n=7) and 66.7% of those with a Master’s degree (n=6) reported some COVID-19 vaccine hesitancy.

One particularly interesting theme that emerged from the interviews was the divergent degrees of health literacy among HCWs when it came to understanding the purpose of vaccination. In this illustrative quote, an ICU/COVID-19-unit nurse explained his perspective that unlike with the polio vaccine decades ago, the COVID-19 vaccine was not a silver bullet guaranteeing complete prevention which is what people were “*accustomed to*” and expected. This then made the decision-making calculus more complex when it came to the COVID-19 vaccine: *“[People] got all the shots like they were supposed to, and they still got COVID… So, that was kind of disconcerting… Symptom reduction is definitely better, but it still is kind of the first of its kind where you can be triple vaxxed, triple boosted, and you still get COVID. And that’s frustrating.” (HCW18)*.

### Structural Dimensions

This study identified factors within the healthcare system itself that impacted HCW vaccine hesitancy towards COVID-19: hospital tier, unit assignment and professional role each influenced what HCWs were exposed to during the pandemic in terms of patient volume, disease severity and poor outcomes. This impacted subsequent decision making regarding COVID-19 vaccination.

#### Hospital Tier and COVID-19 Vaccine Hesitancy

HCWs at JHH, a reference hospital with specialized facilities for managing contagious respiratory pathogens, described encountering a high volume of critically ill COVID-19 patients early in the pandemic when knowledge about transmission and management were limited. JHH HCWs explained that while other hospitals built their capacity overtime, JHH’s rapid response capabilities led to them receiving numerous patient transfers during the initial months of the pandemic. As expressed by one JHH ICU physician, *“It felt like we were kind of designed to do exactly what was needed of us in the initial wave and in subsequent waves, and as a result, I think all the staff were disproportionately exposed to just how awful the disease can be.” (HCW27).* JHH HCWs described how this “*shock*” heightened their awareness of COVID-19 disease severity, personal feelings of vulnerability to illness, and ultimately, eagerness to take the vaccine. This same HCW continued to capture this experience saying,*“ I think seeing your hospital fundamentally change and seeing so many sick patients and personally interacting with so many people who end up dying of COVID… I think it did lead to a lot of people feeling their own vulnerability.”*

#### Hospital Unit, Role and COVID-19 Vaccine Hesitancy

HCWs at both hospitals agreed that relative risk of exposure in different departments was a likely factor in their decisions to get vaccinated. COVID-19 vaccine hesitancy was highest among those who worked outside of the units with the highest rates of severe disease during the pandemic (i.e., ED, ICUs, or COVID-19 units). These HCWs explained that although others may have been physically present in the hospital during the pandemic, they did not necessarily witness the serious nature of the disease firsthand – especially during the initial surge. An ED PA reflected, *“Those were the hardest shifts I’d worked in 20 years. There were more people in the ED than I’d ever seen at one time. Everybody was positive.“(HCW02)*.

Some HCWs recounted that their colleagues who expressed vaccine hesitancy assumed that given their occupational exposure, they had probably already contracted the virus asymptomatically, “*I think they just felt like ‘I’m here way too much. There’s no way I haven’t gotten it somehow*.’,”. They believed they could simply “*rely on [their] ‘innate’ immune system*” for protection from future infection, although this is not the recognized role of the innate immune response.

HCWs at both hospitals also highlighted how their specific roles as nurses, clinical social workers etc. shaped their personal experiences and observations during the pandemic. For many of those in units with the highest rates of disease – where close patient contact was routine – witnessing patients suffering and severe outcomes despite intervention served as a strong motivator to seek vaccination at the first opportunity. As recalled by one ICU/COVID Unit nurse, *“We saw all the intubations. We saw people gasping for air when they were on 100% oxygen. We would be blasting oxygen into their nose and mouth. And they were still gasping for air… So, we wanted to negate that possibility of that happening to us.” (HCW18).* A respiratory therapist echoed these sentiments saying, *“…[we] are the ones who remove people from life support, so my mind was already made up just watching what I had watched. For 22 years it’s been something that I’ve done, and it’s never bothered me, but doing it in such large quantities and watching – literally being the person in the room with families over FaceTime was probably what propelled me the most to make these [COVID-19 vaccination] decisions…” (HCW10)*.

#### Institutional Policies

Additional structural dimensions that impacted HCWs’ decisions to be vaccinated included institutional policies on vaccine mandates. Several HCWs at both hospitals also cited the availability of on-site vaccination as greatly facilitating both influenza and COVID-19 immunization for hospital staff.

##### Johns Hopkins Hospital System Vaccine Mandates

Most HCWs viewed influenza vaccination as a routine job requirement due to the longstanding mandate. They also acknowledged vaccination as part of their professional responsibility to protect patients. An ICU physician explained, *“I think we should be required to do things that put patients in a safer place. We wash our hands. If we can take a vaccine and potentially prevent the spread of the flu to somebody who’s old or immunocompromised, we should do it. It just seems like such a minimal thing.” (HCW23)*. Although some thought that it would have been preferable if a mandate was not necessary, most HCWs interviewed across both hospitals considered the COVID-19 vaccine mandate both foreseeable given trends seen in other states, and “*reasonable*” given the high rates of morbidity and mortality seen in the hospitals.

Numerous HCWs at both hospitals also reported being unaffected by the COVID-19 mandate as they had already decided to vaccinate. Staff supervisors shared that when advising vaccine hesitant staff, they emphasized the mandatory nature of the policy and explained the safety profile of the vaccine and its role in protecting hospital staff, their patients and families.

#### A Pro-vaccine Environment

JHH HCWs perceived low levels of COVID-19 vaccine hesitancy among their colleagues, speculating this might be due to the hospital’s reputation as a leading academic medical center at the forefront of evidence-based medicine. HCWs also noted that Johns Hopkins University is a trusted source of healthcare information, exemplified by initiatives like the widely recognized Johns Hopkins Coronavirus Resource Center[17]. Participants felt that access to cutting-edge research and education within this setting fostered a culture of informed decision-making among staff leading to a group less susceptible to vaccine hesitancy, *“We are literally surrounded by the world’s experts in this area and therefore have been able to go to lectures on Zoom…hear the data straight from the horse’s mouth… I think our community trusts the Johns Hopkins community and knows that these halls are filled with some of the most brilliant minds in the world.” (HCW06).* One JHH ED HCW aptly described the impact of a pro-vaccine environment on personal decision making by borrowing the term “*herd effect*” from vaccine epidemiology, *“If I had worked around 300 coworkers or 500 coworkers that were against [the COVID-19 vaccine], I can’t say how that would have affected my own reservations or hesitation, but I was in the opposite. I was in an environment where we were all kind of wanting that layer of protection.” (HCW02)*. This effect was manifested through peer motivation including initiatives by safety officers, vaccine champions and open discussions with colleagues – particularly those from infection and disease control departments.

### Health Messaging

HCWs evaluated health messages by source, content and mode of delivery, and discussed how these factors impacted vaccine hesitancy.

#### Sources of Information

Findings revealed a complex interplay between different sources of information on HCW vaccination decision making and how each shaped vaccine hesitancy. HCWs reported relying primarily on trusted sources such as medical literature and public health institutions (i.e., CDC, WHO) for information on COVID-19 vaccines. They also reported valuing evidence-based content. News and social media were viewed with skepticism, with most believing they contributed to initial COVID-19 vaccine hesitancy in the general population. Personal providers and colleagues influenced choices to some extent, with friend and family networks playing a mixed role.

##### Workplace Messaging

A recurring theme among HCWs from both hospitals was their reliance on their own institution, Johns Hopkins, as a trusted source of health information throughout the COVID-19 pandemic. Many expressed gratitude for informational meetings with hospital leadership (i.e., Vice President of Medical Affairs, Chief of Infectious Disease) who provided essential updates and clarifications on policy changes. Several JHH HCWs also explained that frequent and regular updates from department and hospital leadership and experts in the form of virtual incident command calls, grand rounds, lectures, townhalls and faculty meetings afforded them opportunities to be well informed about COVID-19 and to ask questions:

> *“[JHH] would have infectious disease experts and vaccine experts and epidemiology experts come. I was getting the top of the world expert medical knowledge on almost a daily basis… I felt like I had the benefit of knowing more than most people in this world, but also, it came from trusted sources. Not news from NBC, CNN, Fox News…I was getting it directly from medical experts, who are also presenting objective data, objective research.” (HCW06)*.

Many HCWs reported that this open communication fostered a sense of trust and transparency, ultimately contributing positively to their willingness to get vaccinated against COVID-19.

#### Content

Analysis identified three key themes as resonating strongly with HCW vaccine acceptance: protection from disease, scientific or public health data and HCWs heightened exposure to pathogens.

##### Protection from Disease

The desire to protect themselves, their colleagues, and ultimately their patients emerged as a primary motivator for HCW vaccination. HCWs recognized their role as guardians of public health and understood the potential consequences of transmitting vaccine-preventable diseases:

> *“[Protecting yourself, your coworkers and your patients], that’s the mission of the hospital… It’s caring for the whole patient. So, if I’m not healthy I can’t care for the patient.” (HCW12)*

> *“Most people in healthcare come from a place of just wanting to help other people, so messaging that focuses on how getting vaccinated would help your patients would be particularly effective.” (HCW27)*

##### Scientific or Public Health Data

HCWs valued evidence-based messaging that clearly presented the science behind vaccine recommendations and included comparative data such as hospitalization rates, morbidity and mortality statistics, and risk-benefit analyses. One HCW in a management position underscored the powerful influence of direct observation and lived experiences in shaping HCW attitudes towards vaccination by noting that their team required minimal persuasion to get vaccinated, *“I’m in a field of critical thinkers in evidence-based medicine. So, all you kind of have to do is present what’s out there and what the literature is showing or what the initial data shows and most of my staff was used to thinking critically and wearing the same hat that we would to recommend to our patients.” (HCW02)*.

##### Exposure to Pathogens

HCWs voiced a clear preference for messaging that recognized the inherent risk associated with their profession. They appreciated communication that recognized their heightened exposure to infectious diseases and directly addressed their concerns about occupational safety. Multiple HCWs also reported preferring messages that linked their concerns about occupational exposure to clear information on safety protocols, access to PPE and vaccines. One ICU/COVID Unit PA stated, *“Anything that acknowledges [we] work very closely with critically ill patients, I think that grabs my attention in the ICU. Especially during COVID, as someone who’s in a COVID unit with that type of exposure.” (HCW05).* This same HCW continued, *“What [JHH] did great was just acknowledging again our fears, so like, ‘We know you’re in these systems. We know you’re constantly exposed. We know your fear of PPE and what you need and require. This is what we’re doing. This is [when] the vaccines will be available. X-Y-Z’.”*

## DISCUSSION

Our findings align with previous research highlighting the multifaceted nature of HCW vaccine hesitancy and support research on the effectiveness of interventions that promote HCW vaccine acceptance including facilitating access, education, reminders, and vaccine mandates [18–21]. However, our study provides deeper insight into how disease perceptions, risk understanding, and knowledge gaps influence HCW vaccine hesitancy. For example, while other studies have found disparate levels of vaccine hesitancy among HCWs with different levels of education[11], this study further illuminated how divergent degrees of understanding about vaccine function contribute to HCW vaccine hesitancy. The assumption that all vaccines prevent infection – when most vaccines provide protection from morbidity and mortality – is a common misunderstanding which held true here as well[22]. This gap in knowledge about the immune response and the primary role of vaccines, illustrates that *even* HCWs do not necessarily understand all the details of the body’s adaptive immune response to vaccines, especially when their focus is clinical practice. Tailoring future interventions to provide clear, science-based information about disease characteristics and the function of vaccines could present a powerful pathway for promoting vaccine acceptance among HCWs.

Our study findings also revealed the significant influence of lived experiences on HCW vaccination decisions. While other studies also found associations between HCW roles and vaccine hesitancy [4,8,9,20], this study provides additional context revealing how direct exposure to seeing the impact of COVID-19 illness and severe disease at large reference hospitals significantly influenced HCWs eagerness to accept vaccination. Findings on how hospital tier, unit assignment, and professional role led to varied decision-making regarding vaccination further emphasizes the need for tailored interventions that consider the specific context and experiences of different HCW sub-groups[18].

### Psychological Determinants of Vaccine Acceptance

Our study also reinforced the importance of psychological determinants in vaccine acceptance found in other studies [23–25]. Confidence, or trust in the safety and efficacy of vaccines, and the health systems and actors that deliver them [24,25] was generally high among participants. Similar to other studies where trust in authorities played an important role [9,23], confidence in institutional leadership also positively influenced vaccine acceptance.

The concept of collective responsibility, or recognition that individual health decisions impact others and a corresponding willingness to protect them through vaccination, was found to be a strong predictor of vaccination behavior in other studies [14,18,23,24] and is particularly relevant for HCWs given their professional duty to protect patients [21]. In our study, HCWs expressed this idea through their desire to protect their families and framing of vaccination as upholding their professional obligation to prioritize patient safety. While some stated a preference for individual choice (low collective responsibility), for most, professional duty superseded this highlighting the complex role of identity in decision making (i.e., personal autonomy vs. professional identity). Messaging that emphasizes HCWs’ professional responsibility could enhance collective responsibility and foster a pro-vaccine workplace.

Calculation, the information-seeking behavior individuals engage in to decide on vaccination is generally positively correlated with vaccine hesitancy [24,25]. Calculation was not however a major contributor to vaccine hesitancy in our study, even among those who expressed vaccine hesitancy, because the abundance of reliable resources promoted by Johns Hopkins reduced the need for independent searching. This communication strategy minimized calculation which can lead to encountering misinformation (a major challenge during the COVID-19 pandemic [26–29]) and ultimately, vaccine hesitancy. Our study identified anxieties among women regarding COVID-19 vaccine safety and potential effects on fertility – a trend also observed elsewhere [30–32]. Given the significant number of women in healthcare, messaging should address concerns specific to this demographic.

Future strategies should emphasize the psychological determinants of vaccine acceptance by building confidence and a sense of collective responsibility, while minimizing constraints and calculation that hinder vaccine acceptance.

### Communication Strategies

Findings also highlight the importance of open communication through various channels to address HCW concerns and suggest that in addition to understanding the “why” behind vaccine hesitancy, strategically designed prompts and reminders can play an important role in encouraging vaccine uptake. The observed pattern of early vaccination among non-hesitant HCWs and delayed vaccination among hesitant individuals suggests the need for flexible campaigns with multi-pronged approaches to information dissemination. Tailored messaging could target early adopters, those who wait-and-see, and the truly hesitant populations with distinct strategies.

### Recommendations

Based on our findings, recommendations for developing vaccine messaging that targets HCWs during novel virus outbreaks are as follows:

- **Recognize that messages will be consumed in their context.** Structural and environmental dimensions can create an enabling and affirming environment that influence employee perceptions of trust and transparency. Having those factors in place can enable an openness to institutional messaging; without established trust, interventions will run up against the limitations of messaging.
- As HCWs seemed to find their personal professional experiences, including their exposure to severe illness, to be the most compelling motivator, **developing content that sets expectations, speaks to the nature of HCW’s jobs and reflects workplace experiences can be a positive driver for vaccine uptake.** A key part of that messaging can include vaccine advocacy from trusted on-the-ground unit leaders with shared levels of exposure and risk.
- HCWs are not a monolith. HCWs from different departments with distinct roles across the hospital system will likely respond to different messaging**. Messages should be tailored for different hospital units and HCW roles.** Hospitals also need to think about messages and modes of delivery that might be more effective for micro-targeting groups at different points in time, especially during an evolving event.

### Limitations

This study has several limitations that may impact how broadly its findings can be applied. Since all interviews occurred after the emergence of COVID-19, HCWs’ perspectives on past experiences with influenza and vaccination may have been influenced by the heightened focus on infectious diseases. Additionally, vaccination decisions can change over time; this study captures a snapshot of HCW attitudes during data collection in 2022. The focus on a single healthcare system and the smaller sample size of qualitative research also limits generalizability to other settings, regions, and HCW populations with different characteristics. Selection bias is also a concern; volunteers may have had stronger opinions on vaccination and only one key informant was interviewed, potentially skewing results. All participants were fully vaccinated, so their reasons for getting vaccinated may differ from their unvaccinated peers. Finally, efforts to recruit more minorities were unsuccessful, limiting participant diversity – likely an important source of bias. These limitations underscore the need for further research on HCWs’ perspectives on vaccination across a wider range of settings and populations.

### Conclusion

HCWs present a nuanced challenge in vaccine hesitancy research. With their extensive healthcare knowledge and history of employment which mandates vaccination, broad national strategies targeting vaccine-hesitant individuals do not translate effectively to this population. Understanding how to improve vaccine acceptance among HCWs requires understanding more than the simplistic binaries of measuring vaccine acceptance or refusal. Our study explores the interplay of individual (i.e., perceptions of disease severity and susceptibility), structural (i.e., institutional policies and health system factors), and environmental (i.e., peer networks and transparent communication) dynamics influencing HCW decision-making. These findings have important public health implications and provide insights for designing and delivering targeted vaccination campaigns tailored to HCWs facing heightened occupational exposure to viral respiratory illnesses – particularly in the context of a novel virus outbreak.

## Data Availability

All data produced in the present work are contained in the manuscript.

## ACKNOWLEDGEMENTS

The authors would like to acknowledge the National Institutes of Health (NIH) National Institute of Allergy and Infectious Disease (NIAID) for funding this research, and thank Wendy Bennett, Katherine Fenstermacher, Patrick Shea and Anna Yin for their support, and the healthcare professionals of the Johns Hopkins Health System for their participation in this study. Preliminary study findings were presented in Washington, DC at the Association of American Medical Colleges (AAMC) Health Workforce Research Conference (May 2023) and Baltimore, MD at the 2nd Centers of Excellence for Influenza Research and Response (CEIRR) Network Annual Network Meeting (August 2023).

## CONTRIBUTIONS

Funding acquisition: ENR, SK, AP, RM. Study conceptualization, design and methodology: ENR, AP, RM. Recruitment, data collection and analysis: ENR. Original manuscript draft preparation and writing: ENR. Manuscript review and editing. ENR, SK, RER, AP, RM.

All authors read and approved the final manuscript.

## REFERENCES

1. Gómez-Ochoa SA, Franco OH, Rojas LZ, Raguindin PF, Roa-Díaz ZM, Wyssmann BM, et al. COVID-19 in Health-Care Workers: A Living Systematic Review and Meta-Analysis of Prevalence, Risk Factors, Clinical Characteristics, and Outcomes. Am J Epidemiol [Internet]. 2021 [cited 2023 Dec 3];190:161–75. Available from: 10.1093/aje/kwaa191

2. Nguyen LH, Drew DA, Graham MS, Joshi AD, Guo CG, Ma W, et al. Risk of COVID-19 among front-line health-care workers and the general community: a prospective cohort study. Lancet Public Health. 2020;5:e475–83.

3. Vilches TN, Sah P, Moghadas SM, Shoukat A, Fitzpatrick MC, Hotez PJ, et al. COVID-19 hospitalizations and deaths averted under an accelerated vaccination program in northeastern and southern regions of the USA. The Lancet Regional Health - Americas [Internet]. 2022;6:100147. Available from: 10.1016/j.

4. Peterson CJ, Lee B, Nugent K. COVID-19 Vaccination Hesitancy among Healthcare Workers—A Review. Vaccines (Basel). MDPI; 2022.

5. World Health Organization. Strategy to Achieve Global Covid-19 Vaccination by mid-2022 [Internet]. Geneva; 2021 Oct. Available from: https://www.who.int/publications/m/item/strategy-to-achieve-global-covid-19-vaccination-by-mid-2022

6. Dooling K, McClung N, Chamberland M, Marin M, Wallace M, Bell B, et al. The Advisory Committee on Immunization Practices’ Interim Recommendation for Allocating Initial Supplies of COVID-19 Vaccine — United States, 2020. Morbidity and Mortality Weekly Report. 2020;69:1857–9.

7. MacDonald NE, Eskola J, Liang X, Chaudhuri M, Dube E, Gellin B, et al. Vaccine hesitancy: Definition, scope and determinants. Vaccine [Internet]. 2015 [cited 2023 Jan 28];33:4161–4. Available from: https://pubmed.ncbi.nlm.nih.gov/25896383/

8. Liz Hamel, Ashley Kirzinger, Cailey Muñana, Mollyann Brodie. KFF COVID-19 Vaccine Monitor: December 2020 | KFF [Internet]. 2020 [cited 2023 Jan 9]. Available from: https://www.kff.org/coronavirus-covid-19/report/kff-covid-19-vaccine-monitor-december-2020/

9. Shaw J, Stewart T, Anderson KB, Hanley S, Thomas SJ, Salmon DA, et al. Assessment of US Healthcare Personnel Attitudes Towards Coronavirus Disease 2019 (COVID-19) Vaccination in a Large University Healthcare System. Clinical Infectious Diseases [Internet]. 2021 [cited 2023 Mar 29];73:1776–83. Available from: https://academic.oup.com/cid/article/73/10/1776/6118651

10. Brita Roy, Vineet Kumar, Arjun Venkatesh. Health Care Workers’ Reluctance to Take the Covid-19 Vaccine: A Consumer-Marketing Approach to Identifying and Overcoming Hesitancy. NEJM Catal [Internet]. 2020 [cited 2023 Dec 3]; Available from: https://catalyst.nejm.org/doi/full/10.1056/CAT.20.0676

11. Lazer D, Ognyanova K, Green J, Baum M, Druckman J, Gitomer A, et al. The COVID States Project #47: Update on COVID-19 vaccine attitudes among healthcare workers [Internet]. OSF Preprints; 2021 Mar. Available from: https://osf.io/a352z/

12. Baniak LM, Luyster FS, Raible CA, McCray EE, Strollo PJ. COVID-19 Vaccine Hesitancy and Uptake among Nursing Staff during an Active Vaccine Rollout. Vaccines (Basel). 2021;9.

13. Browne S, Feemster K, Shen A, Green-McKenzie J, Momplaisir F, Faig W, et al. Covid-19 vaccine hesitancy among physicians, physician assistants, nurse practitioners, and nurses in two academic hospitals in Philadelphia. Infect Control Hosp Epidemiol. 2021;1–24.

14. Kwok KO, Li KK, WEI WI, Tang A, Wong SYS, Lee SS. Influenza vaccine uptake, COVID-19 vaccination intention and vaccine hesitancy among nurses: A survey. Int J Nurs Stud. 2021;114.

15. Johns Hopkins Medicine. The Johns Hopkins Health System Corporation [Internet]. About Johns Hopkins Medicine. 2024 [cited 2024 May 31]. Available from: https://www.hopkinsmedicine.org/about/johnshopkins-health-system-corp

16. Hennink M, Kaiser BN. Sample sizes for saturation in qualitative research: A systematic review of empirical tests. Soc Sci Med. 2022;292.

17. Johns Hopkins Coronavirus Resource Center [Internet]. Johns Hopkins University & Medicine. [cited 2024 Feb 10]. Available from: https://coronavirus.jhu.edu/

18. Thaivalappil A, Young I, MacKay M, Pearl DL, Papadopoulos A. A qualitative study exploring healthcare providers’ and trainees’ barriers to COVID-19 and influenza vaccine uptake. Health Psychol Behav Med. 2022;10:695–712.

19. Hollmeyer H, Hayden F, Mounts A, Buchholz U. Review: Interventions to increase influenza vaccination among healthcare workers in hospitals. Influenza Other Respir Viruses. 2013;7:604–21.

20. Kandiah S, Iheaku O, Farrque M, Hanna J, Johnson KB, Wiley Z, et al. COVID-19 Vaccine Hesitancy Among Health Care Workers in Four Health Care Systems in Atlanta. Open Forum Infect Dis. 2022;9.

21. Gur-Arie R, Berger Z, Reiss DR. COVID-19 Vaccine Uptake Through the Lived Experiences of Health Care Personnel: Policy and Legal Considerations. Health Equity. 2021;5:688–96.

22. Tentori K, Passerini A, Timberlake B, Pighin S. The misunderstanding of vaccine efficacy. Soc Sci Med. 2021;289.

23. Wismans A, Thurik R, Baptista R, Dejardin M, Janssen F, Franken I. Psychological characteristics and the mediating role of the 5C Model in explaining students’ COVID-19 vaccination intention. PLoS One. 2021;16.

24. Betsch C, Schmid P, Heinemeier D, Korn L, Holtmann C, Böhm R. Beyond confidence: Development of a measure assessing the 5C psychological antecedents of vaccination. PLoS One. 2018;13.

25. Betsch C, Bach Habersaat K, Deshevoi S, Heinemeier D, Briko N, Kostenko N, et al. Sample study protocol for adapting and translating the 5C scale to assess the psychological antecedents of vaccination. BMJ Open. 2020;10.

26. World Health Organization. Fighting Misinformation in the Time of COVID-19, One Click at a Time. WHO Newsroom. 2021.

27. Hwang J, Borah P, Shah D, Brauer M. The relationship among covid-19 information seeking, news media use, and emotional distress at the onset of the pandemic. Int J Environ Res Public Health. 2021;18.

28. Caceres MMF, Sosa JP, Lawrence JA, Sestacovschi C, Tidd-Johnson A, UI Rasool MH, et al. The impact of misinformation on the COVID-19 pandemic. AIMS Public Health. American Institute of Mathematical Sciences; 2022. p. 262–77.

29. Malik A, Bashir F, Mahmood K. Antecedents and Consequences of Misinformation Sharing Behavior among Adults on Social Media during COVID-19. Sage Open. 2023;13.

30. Cox E, Sanchez M, Baxter C, Crary I, Every E, Munson J, et al. COVID-19 Vaccine Hesitancy among English-Speaking Pregnant Women Living in Rural Western United States. Vaccines (Basel). 2023;11.

31. Shamshirsaz AA, Hessami K, Morain S, Afshar Y, Nassr AA, Arian SE, et al. Intention to Receive COVID-19 Vaccine during Pregnancy: A Systematic Review and Meta-analysis. Am J Perinatol. Thieme Medical Publishers, Inc.; 2022. p. 492–500.

32. Rawal S, Tackett RL, Stone RH, Young HN. COVID-19 vaccination among pregnant people in the United States: a systematic review. 2022; Available from: 10.1016/j.

